# Association between functional ability, chronic diseases and lifestyle risk factors in older community dwelling adults: protocol for a prospective observational Chinese cohort study

**DOI:** 10.1101/2024.04.22.24306157

**Authors:** Xin Wei, Xu Dan, Dou Zulin, Jacques Angela, Umbella Josephine, Fan Yuling, Zhang Longsheng, Yang Haiwen, Cai Hong, Hill Anne-Marie

**Author notes:** ORCID IDs Wei Xin https://orcid.org/0000-0002-3009-9406 Anne-Marie Hill http://orcid.org/0000-0003-1411-6752 Zulin Dou https://orcid.org/0000-0001-8904-6440 Dan Xu http://orcid.org/0000-0001-6649-1111 Josephine Umbella https://orcid.org/0000-0001-5636-4565 Angela Jacques http://orcid.org/0000-0002-0461-681X.

## Abstract

**Background:** The increasing proportion of the ageing population has become a significant focus for healthcare and social services in many countries worldwide, including China. Impaired physical fitness or the presence of chronic diseases may decrease functional ability and health related quality of life for older adults. The aim of the study is to investigate the association between functional abilities, including motor and cognitive function, with lifestyle risk factors, including activities of daily living (ADL), physical activity falls, frailty, and chronic diseases in older adults living in Guangzhou, China.

**Methods:** A prospective observational cohort study will be conducted. Older adults aged 60 years and over living in urban Guangzhou, China will be eligible for inclusion. This study will be conducted in a community health service centre in Guangzhou. Inclusion criteria are that older adults can independently ambulate short distances indoors with or without a walking aid and can provide written informed consent. The outcomes are functional ability measured using, i Timed Up and Go test (TUGT), gait speed, handgrip strength and Functional Reach Test (FRT), cognition measured using the Mini-Cog, ADL measured with the Lawton Instrumental Activities of Daily Living Scale (Lawton IADL), HR-QoL measured using the European Quality of Life 5 Dimensions 3 Level Version (EQ5D3L), physical activity, falls history, lifestyle risk factors, anthropometric data, presence of chronic diseases measured by standardized blood tests, and blood pressure. Data collection will be performed by two senior physiotherapists and nurses working in the community centre.

**Trial registration:** Ethical approval (II2023-155-02) for this study was obtained from the Health Research Ethics Committee of the Third Affiliated Hospital of Sun Yat-Sen University, Guangzhou, China.

**Chinese Clinical Trial Registry Centre:** ChiCTR2300076095.

## Background

The proportion of older adults in the population is increasing worldwide and by 2050 22.0% of total population will be aged 60 years or over ^1^. The increased ageing population necessitates changes for healthcare and social services in national health systems ^2^ ^3^. The increasing ageing population has unique regional specificity, particularly in China, the largest developing country in Asia ^3^. As of 2022, the total population of mainland China is 1.4 billion, with the number of individuals aged 65 and above exceeding 164.5 million ^2^. By 2050, China’s ageing population is projected to increase sharply to 400 million, accounting for 30% of the total population, positioning China at the forefront of the global ageing trend ^2^.

Functional ability is defined as actual or potential capacity of an individual to perform the activities and tasks that can be normally expected, including motor and cognitive functions ^4–7^. Impaired functional ability is associated with loss of independence, which is a key factor in determining an older adult’s health-related quality of life (HR-QoL) ^8^. A cross-sectional Chinese study reported that 26.57% of older adults aged 60 years and over have motor dysfunction and 15.39% have cognitive dysfunction. These problems in functional ability are associated with increases in morbidity and mortality rates, as well as impaired activities of daily living ^9^.

The etiology of decline in physical fitness in older adults is complex, and can be as a result of physiological, neurological, lifestyle, and psychological factors, which may lead to chronic disease ^10^. Firstly, ageing-induced alterations in central and peripheral nervous systems including neuromuscular junction, lead to balance and gait disorders, coordination difficulties, and reduced muscle strength with subsequent high fall risk ^10,11^. High falls prevalence has been identified in recent findings showing that 20-30% of older adults experience falls resulting in mild-to-severe injuries, and more than 50% of injurious falls require inpatient treatment ^8^. This age-induced motor dysfunction and potential consequences of falls are associated with older adults’ loss of independence, restricting their functional abilities and reduced HR-QoL ^12^.

Cognitive function declines with age due to reduced grey matter volume, functional brain changes, associated chronic diseases, and lifestyle risk factors ^13^. A recent systematic review included 80 studies and concluded that the median prevalence of cognitive impairment in older adults was 19.0% and the median incidence was 53.97 per 1000 person-year ^14^. Declined cognitive function in older adults includes memory, sensory perception, attention, and language with potentially compromising functional independence, social skills, increasing health risks and comorbidities, and significantly decreasing quality of life ^13^.

In addition to the ageing process, chronic diseases and lifestyle factors may also contribute to motor and cognitive decline ^15^. For example, people who have diabetes, cancer, and obesity and those who have low levels of physical activity are reported have impairments in functional ability compared to those who exercise or do not have such chronic diseases ^16^. The four most common chronic diseases in Chinese populations which are 60% for hypertension, 51% for hypercholesterolemia, 35% for arthritis, and 29% for ischemic or coronary heart disease ^17^. Differences in the prevalence of chronic diseases between countries may be attributed in part to country-specific lifestyle risk factors, culture, and dietary habits ^18^.

A recent study showed that chronic diseases may contribute to functional decline, falls, frailty and balance impairment in older adults ^19^. Moreover, systematic reviews have reported that cardiovascular risk factors, including diabetes, obesity, and hypertension, are associated with an increased risk of dementia and functional disability in the ageing population ^20–22^.

Lifestyle risk factors, such as physical inactivity, smoking, and alcohol consumption, are associated with chronic diseases, potentially accelerating declines in motor and cognitive function ^23^. Functional decline is also a harbinger of various neurological disorders, including dementia, Parkinson’s disease, and stroke ^24^. Therefore, when investigating how to prevent the onset of chronic diseases, it is important to understand the impact of lifestyle risk factors, and motor and cognitive function on the development of chronic diseases ^19^.

However, although chronic disease is known to be associated with age-related problems, limited studies have investigated the associations between functional abilities, chronic diseases and lifestyle risk factors among older Chinese community-dwelling adults. Our previous systematic review found that community-based rehabilitation can significantly improve balance and walking abilities and handgrip strength for older community-dwelling adults living in Asian countries. However, these studies did not evaluate the presence of chronic diseases and only limited research was conducted in China ^25^. A Chinese self-reported survey reported that while 11.1% of older adults have been referred to CBR services, less than 1% of older adults actually receive these services ^26^. Therefore, the aims of the study are: i) to investigate the association between functional ability, chronic diseases and lifestyle risk factors in older community-dwelling adults living in Guangzhou, China; ii) to conduct a comprehensive needs analysis for CBR service in the population attending the community health service centre in Tianhe community.

## Methods

### Design

This study is a prospective observational study, designed and reported in accordance with the Strengthening the Reporting of Observational Studies in Epidemiology (STROBE) guidelines ^27^.

### Ethical considerations and trial registration

Ethical approval (II2023-155-02) for this study was obtained from the Health Research Ethics Committee of the Third Affiliated Hospital of Sun Yat-Sen University, Guangzhou, China. The study is registered on the Chinese Clinical Trial Registry Centre (registry number: ChiCTR2300076095). All participants will provide written informed consent.

### Participants and Setting

The setting is the Tianhe community health service centre (CHSC) in Guangzhou city, Guangdong province, China. Residents living in the community aged 60 years and over who are registered with the Tianhe CHSC and have undertaken the free health examinations provided by Chinese government, are the population of interest. The MeSH definition of older adults is 65 years and over. However, this study will include adults aged 60 years and over to provide data relevant for the ageing population in China. The Chinese government provides services that focus on preventive health care for this increasing cohort of older adults, to mitigate effects of ageing.

### Inclusion and exclusion criteria

Older adults will be eligible for inclusion in the study if they are able to independently ambulate short distances indoors with or without a walking aid and can provide written informed consent. Older adults will be excluded if they live in a nursing home, are unable to walk without the assistance of another person, are deemed unsuitable by their general practitioner (GP) due to unstable medical conditions, have been diagnosed with a heart attack in the previous three months, have a diagnosis of dementia or are unable to follow instructions or answer the interview questions.

### Recruitment and data collection procedure

Enrolment flyers will be placed at the entrance of the health examination room in the Tianhe CHSC, and an advertisement will be posted on the internet, including WeChat, a popular Chinese social media platform. Additionally, nurses working in the Tianhe CHSC health examination centre will present information about the study to older adults who potentially meet the inclusion criteria. Recruitment and outcome data collection will be conducted by two senior, experienced physiotherapists (WX and XF), who will complete 1.5 hours’ training together before data collection to reduce assessment bias ^28^. Participants will be consecutively recruited if they meet the inclusion and exclusion criteria ^28^.

### Sample size

A sample that represents the broader older population in China will be sought. Annual falls prevalence in the Chinese older adult population has been found to be approximately 23.3% ^29^. The minimum sample size needed to represent a population falls prevalence of 20% at a 5% level of significance ^30^ is n=246 patients. Therefore, this study will aim to enrol n=260 participants (allowing for a dropout rate of approximately 5%).

## Outcomes

### Demographic and physical activity data

Participant information will be collected by conducting face-to-face interviews during their attendance for their annual health examination at the Tianhe CHSC. A questionnaire will be administered to gather demographic data, including age, gender, educational attainment (middle school or lower, high school or university), occupation (physical (such as working in manufacturing settings) or non-physical (such as office based occupations), living situation (lives alone, with family, with partner), smoking status (yes or no (have quit)), alcohol consumption (yes or no (have quit)), number of medicines prescribed, number of falls in the previous year, and report of musculoskeletal pain (including location of pain, e.g. lumbar spine). Physical activity data will be collected using the Physical Activity Scale for the Elderly (PACE) questionnaire, which is a widely used in epidemiological studies and validated for older Chinese adults ^31^.

### Consumer survey of community-based rehabilitation services

Participants will be administered a questionnaire seeking feedback about their experiences and needs regarding CBR services. Questions will include the frequency of CBR services utilised per week, satisfaction rating for CBR services experienced (0-10), and preferences for CBR services to be provided by the CHSC in the future.

### Anthropometric data, blood tests, and blood pressure

Anthropometric data (weight, height, and waist circumference), and blood biochemical examination (including fasting blood glucose, uric acid, high-density lipoprotein, low-density lipoprotein triglyceride, and total cholesterol) will be collected in the morning before breakfast, twice. Blood pressure on the right side will be collected twice after a 5-minute rest, with a one-minute interval between the two measurements. These data will be collected by trained, experienced nurses during the free annual health examination.

### Assessment of motor function, cognitive function, frailty, ADL, and health-related QoL

Physical fitness assessments will be measured twice by the physiotherapists (WX and XF), and the average value of the measurements will be calculated and recorded.

### Motor function

Handgrip strength for both hands will be measured using an electronic handgrip meter (CAMRY, EH101, China). Participants will stand with feet shoulder-width apart, body upright, holding the electronic handgrip meter down, then according to researcher’s instruction, they will exert maximum force for 2 seconds. Adequate rest will be given between the two measurements ^32^. Overall functional ability will be measured by the Timed Up and Go test (TUGT) ^33^, which requires an armless chair and a stopwatch. The TUGT assesses the time taken for participants to stand from the chair, walk 3 meters, turn around, walk back, and sit down. Assessors will walk beside participants during the test to reduce fall risk, without touching or supporting ^34^. Gait speed will be measured by a 5-meter walking test, with 2 meters for acceleration and deceleration, asking participants to walk at a self-selected speed ^35^. Dynamic balance will be measured by the Functional Reach Test (FRT), where participants stand next to a wall, fist closed, shoulder flexed at 90 degrees, then reach forward as far as possible without stepping. The distance between the start and end positions will be recorded (cm) ^36^. Frailty will be assessed using the FRAIL scale, which includes questions on fatigue, resistance, ambulation, illness, and weight loss ^19^. The score ranges from 0 to 5 and a score of 3-5 indicates frailty, 1 to 2 indicates prefrailty, and 0 represents robust health status.

### Cognitive function

Cognition will be assessed using the Mini-Cog test, which combines a delayed-recall (DR) test and a clock-drawing test (CDT), with a total score of 5 ^37^. Participants who score 3, 4 or 5 will be recorded as “No”, which indicates lower likelihood of dementia. By contrast, a score of 0, 1, or 2 indicates a higher likelihood of clinically important cognitive dysfunction and will be recorded as “Yes.” These participants will then be referred to a general practitioner s for further dementia assessments, such as a mini-mental state examination ^37^. If the participants are diagnosed by their GP as having dementia they will be excluded.

### ADL and health related HR-QoL

Lawton Instrumental Activities of Daily Living Scale (Lawton IADL) will be used to assess instrumental activities of daily living. The Lawton IADL scale contains 8 items: use of telephone, transportation, shopping, meal preparation, housework, laundry, medication management, and managing finances, with the total score ranging from 0 (low function) to 8 (high function) ^38^. This scale will be administered to participants through a face-to-face interview during the data collection procedure. HR-QoL will be measured using the European Quality of Life 5 Dimensions 3 Level Version (EQ5D3L), a patient-reported outcomes measurement ^39^. The five dimensions include mobility, self-care, usual activities, pain/discomfort, and anxiety/depression, each with three rating levels (no problems, some problems, and extreme problems). An EQ visual analogue scale (VAS) score is also collected. This scale consists of a vertical 100-point visual analogue scale, that asks participants for a self-reported health rating at the time of assessment, with 100 points representing the best possible health status.

## Chronic diseases

The presence of up to six chronic diseases will be recorded for each participant by the two physiotherapists. Diagnosis of chronic diseases will be determined by the CBR centre treating doctor. Diagnosis is determined according to the following criteria:

(1) Hypertension: systolic blood pressure on (SBP) left arm ≥ 130 mm Hg ^40^.
(2) Hyperuricemia: uricemia (UA), indicating impaired kidney function: male >420 μmol/L, female >350 μmol/L ^41^.
(3) Hyperglycemia (diabetes): fasting blood glucose (FBG) ≥ 7.0 mmol/L; based on the WHO diagnostic criteria for diabetes ^42^.
(4) Hypercholesterolemia: triglyceride (TG) ≥2.3mmol/L or total cholesterol ≥6.2mmol/L (TC); based on the guidelines for the prevention and treatment of dyslipidemia or adults in China ^42^.
(5) Dyslipidemia: low density lipoprotein (LDL) ≥4.1mmol/L or high-density lipoprotein (HDL) <1.0 mmol/L ^42^.
(6) Overweight: 28 kg/m2>body mass index (BMI)≥24 kg/m2, Obesity: ≥28 kg/m2 ^41^.

## Statistical analysis

Data will be summarised using descriptive statistics including means and standard deviations or medians and interquartile ranges for continuous data and frequency distributions for categorical data. Univariate group comparisons of health related functional and QOL outcomes between age and gender groups will be performed using Mann-Whitney U or t tests for continuous data and χ2 or Fisher’s exact tests for categorical data. Associations between outcomes (including TUG score, gait speed, handgrip strength and FRT distance) and patient characteristics (including age, gender, amount of physical activity, anthropometric and sociodemographic factors, and chronic diseases) will be examined using generalised linear regression models, adjusting for covariates known to be associated with outcomes. Results will be summarised using beta coefficients and 95% confidence intervals. All hypotheses will be two-tailed and significance levels set at alpha=0.05. Stata version 18 (StataCorp, College Station, TX) will be used for data analysis.

## Discussion

Findings from this study will be useful to inform the development of community healthcare services for older adults in developing countries. The study will also provide evidence-based knowledge for clinical practitioners regarding the risk factors for decline in functional abilities among older community-dwelling adults living with chronic diseases.

Findings informed by local cultural contexts can aid clinicians in providing efficacious rehabilitation interventions for community-dwelling older adults with chronic diseases. The study can also guide the tailoring of public health policies and the refinement of healthcare delivery systems for ageing-related chronic diseases.

Therefore, the results of this study will address a critical gap in geriatric healthcare, in the development of CBR services, potentially guiding the refinement of CBR programs in China and comparable developing countries that also have ageing populations.

Strengths of the study are that it will employ a prospective design, according to a published protocol, and therefore provides a higher level of evidence than a retrospective cohort. The study will conduct a comprehensive assessment of multiple functional outcomes using trained physiotherapists and chronic diseases are accurately diagnosed using blood test results. This increases the reliability of the findings and their generalisability to older populations with chronic disease. The study design is informed by, and reported according to the STROBE guidelines, to reduce the method bias. Motor and cognitive function and physical activity outcomes will be measured utilising reliable and valid assessments in China for to increase the reliability of the results.

This study has limitations. It will be conducted at a single centre, which may reduce the generalisability of the results. Only older adults who can ambulate at least short distances independently will be included. Therefore, specific findings and recommendations about how older community dwelling adults who have disability and are unable walk unaided cannot be made to Chinese CBR centres.

This protocol reports the methods for a prospective cohort study in one city in China, a developing country that has an ageing population. Further longitudinal studies that explore the long-term effects of chronic diseases on functional ability are required. These should include multiple CBR centres in developing countries.

## Competing interests

The authors declare that they have no conflict of interest.

## Funding

This study is funded by the National Key Research &Development Program of China (Grant No. 2020YFC2004205). Anne-Marie Hill is supported by a National Health and Medical Research Council (NHMRC) of Australia Investigator (EL2) award (GNT1174179) and the Royal Perth Hospital Research Foundation. Wei Xin is supported by a Curtin University Fee Offset Scholarship for her PhD.

## Authors’ contributions

WX was primarily responsible for drafting the manuscript with support from AMH and DX. WX, AMH, DX and ZD were responsible for study conception and design with support from AJ. AJ, AMH and WX completed the statistical analyses. WX and UA performed data translation. YF, LZ, HY, and HC contribute to ethics procedures, study procedure, data collection and data management at the sites. All authors critically reviewed the manuscript for its content and approved the final version of the manuscript for submission.

## Data Availability

All data produced in the present study are available upon reasonable request to the authors

## Acknowledgements

We are grateful for the support provided by the community health service centre.

## Notes

### Competing Interest Statement

The authors have declared no competing interest.

### Clinical Trial

ChiCTR2300076095

### Author Declarations

Trial registration: Ethical approval (II2023-155-02) for this study was obtained from the Health Research Ethics Committee of the Third Affiliated Hospital of Sun Yat-Sen University, Guangzhou, China. Chinese Clinical Trial Registry Centre: ChiCTR2300076095

